# Qualitative analysis of the Life Skills training strategy at a public university in Colombia, 2024

**DOI:** 10.64898/2026.07.16.26358291

**Authors:** Nicolas Ortiz Ruiz, Yamileth López Paz, Yenifer Patricia Orobio Lerma, Delia Burgos Dávila, Heidy Johana Medina Zapata, Katherine Paola Manzano Valencia, Alexander Almeida Espinosa

## Abstract

This qualitative study evaluated the Life Skills (HpV) training strategy at a public university in Colombia in 2024, analyzing its impact on students’ positive mental health and psychosocial competencies. The mixed-methods research employed semi-structured interviews with faculty and three student focus groups, using thematic analysis to categorize strengths, weaknesses, and perceived changes. Results highlighted curricular coherence, academic freedom, and participatory methodology as key aspects, fostering self-awareness, emotional management, and the building of support networks. Students reported improvements in well-being, stress management, and academic performance, though challenges such as initial resistance to emotional content and student diversity were identified.

The conclusions underscore the value of experiential courses in university education, promoting horizontal relationships and safe spaces for collective reflection. Future studies are recommended to expand participant diversity and incorporate quantitative data triangulation to further explore the intervention’s effects.

## Introduction

In the university context, student well-being and educational experience are affected by various psychosocial issues, including family conflicts, feelings of loneliness, low self-esteem, emotional imbalances, self-care difficulties, problematic substance use, and conflictive interpersonal relationships [1–3]. Research indicates that factors such as adverse socioeconomic conditions, high parental expectations, and an educational model centered on productivity generate emotional tensions [4–5]. These situations negatively impact academic performance, mental health, and student retention, highlighting the critical need to strengthen coping mechanisms, emotional management, and support networks [6].

Despite structural constraints, student agency allows individuals to reflect and act within their contexts, thereby influencing their educational trajectories, mental health, and social mobility [7]. Consequently, it is essential to promote an education that fosters skills for building meaningful relationships and transforming environments through both individual and collective care [8].

This necessitates comprehensive approaches that address the individual, social, and cultural dimensions of Positive Mental Health (PMH). Such approaches enable students to navigate psychological distress—defined as a form of malaise not reducible to diagnostic categories but related to subjective experiences and environmental links, often associated with intense emotions such as sadness, anxiety, fear, anger, or guilt [9–10]. The PMH framework encompasses dimensions such as self-concept, self-esteem, emotional self-regulation, empathy, and social skills required to face daily vicissitudes and psychological distress [11–12]. Furthermore, the World Health Organization and the Pan American Health Organization promote Life Skills (LS) as psychosocial capacities essential for managing daily challenges and establishing healthy relationships, which are fundamental for emotional well-being and risk prevention [13–14].

Universities have implemented socio- and psycho-educational strategies to promote mental health, focusing on preventing emotional distress, reducing the stigma associated with mental disorders, and strengthening socio-emotional skills [1,3,5,15–16]. Programs in Spanish and Latin American universities have yielded positive results in transforming attitudes toward mental health and increasing the willingness to seek help [17]. Additionally, interventions such as mindfulness-based therapy have been shown to reduce symptoms of depression, anxiety, and stress while enhancing emotional regulation [18]. However, some critics warn against individualizing approaches that may overlook the structural conditions underlying psychological malaise [19].

Higher education institutions face the challenge of integrating emotional and ethical development into their curricula, strengthening personal growth courses, and revising curricular designs to account for student diversity [5,16,20]. Participation and listening spaces are vital for fostering a sense of belonging and well-being [5–6,8]. In Colombia, the Department for Social Prosperity (DPS) integrates LS training for participants in programs such as *Jóvenes en Acción* and *Familias en Acción* to complement formal education and facilitate successful transitions toward employment and entrepreneurship [21].

Specifically, the LS course implemented at a public university in Colombia—the focus of this analysis—serves as a strategy for integral formation, strengthening students’ emotional, communicative, and ethical competencies. Since its implementation in 2018, the course has aligned with institutional policies promoting academic flexibility, critical thinking, and ethical responsibility. It aims to foster well-being and PMH by articulating with institutional pillars such as University Well-being, Integral Formation, Knowledge Generation, and Social Engagement.

This article presents the qualitative results of a mixed-methods study that evaluated the LS training strategy at two campuses of a Colombian public university in 2024. The research identified changes in life skills and PMH factors, assessed the alignment of the curricular design, and examined the successes and challenges from the perspectives of both faculty and students. This paper focuses primarily on the latter objective, relating it to the skill transformations reported by participants. The qualitative findings aim to provide actionable knowledge to strengthen the strategy and improve institutional conditions for a university education centered on care, well-being, and social transformation. This research was conducted by two institutional research groups and was supported by internal funding.

## Materials and Methods

This article presents the results of a mixed-methods, quasi-experimental, longitudinal, and prospective evaluative study. The research was approved by the Institutional Ethics Review Committee for Research with Human Beings of the participating institution, with approval act 013-024 of 2024 and was carried out on two campuses of a public university in Colombia.

The evaluated intervention is the course titled “Life Skills: Discovering the emotional, affective, cognitive, and social self,” which aims to contribute to human development, well-being, and quality of life. The curriculum seeks to balance humanistic, scientific, and technological dimensions while stimulating individual, social, and professional growth. The course is delivered through 16 three-hour in-person workshop sessions. These sessions address core competencies, including self-awareness, empathy, assertive communication, stress management, emotional regulation, interpersonal relationships, self-esteem, decision-making, conflict resolution, critical and creative thinking, and the prevention of psychoactive substance use. The pedagogical model employs participatory methodologies centered on reflective and experiential dialogue to foster autonomy, critical analysis, and collective motivation.

In the quantitative component, standardized instruments validated for university settings were administered to assess Life Skills (LS) and Positive Mental Health (PMH). A pre-test and post-test design was applied to a non-probability sample of 298 students. Data analysis included frequency and percentage tables, as well as the measurement of concordance between LS and PMH scores to identify significant intervention-driven changes.

Regarding the qualitative component, a non-probability convenience sampling method was utilized [22]. Semi-structured interviews were conducted with the total faculty population (seven instructors) who taught the course during the second semester of 2024. Additionally, three focus groups (one at each campus) were held with 24 students ranging from the first to the tenth academic semester. Written informed consent was obtained from all participating teachers and students after the ethics committee approved this consent.

Data were recorded and transcribed verbatim, then processed using Excel® matrices. Subsequently, a thematic content analysis was performed, focusing on the following categories: the reliability of the course design, its implementation, and identified strengths and weaknesses [23].

**Table 1.** Academic Background of Participating Faculty.

| Undergraduate Degree | Graduate Degree |
| --- | --- |
| Four Social Workers | ● Two MSc in Public Health<br>● One MSc in Psychology<br>● One MA in Social Intervention |
| BA in Social Sciences | MA in Urban Studies |
| Sociologist | MA in Anthropology |
| Physician | MSc in Mental Health |

*Source: Prepared by the authors*

## Results

### The main research findings are presented below, starting with the strengths, followed by the challenges

The first strength of the course lies in the existence of a solid and coherent syllabus, which is considered a key element of the curriculum. The interviewed instructors speak positively of how the learning outcomes, clearly defined topics (LS), supporting written and audiovisual materials, and methodological tools are integrated. They highlight that, although each topic is distinct, they are interconnected, allowing for linkages and cumulative progress throughout the course that reflects the combinations and the complex, multidimensional nature of LS. Regarding this point, one of the interviewees states:

> *“I find that very coherent. Furthermore, one of the great things about the subject matter is that I can talk about relationships in the third class, but also in the fourth—and in the fourth, I can talk about communication—so they all intertwine and are completely connected” (Teacher 1)*.

They agree in identifying academic freedom as a strength; although there is an established syllabus serving as a common foundation, each instructor is free to adapt the course’s progression based on the group’s characteristics and needs, as well as the atmosphere that evolves throughout the term. This lends an emergent quality to the curriculum and fosters a dynamic of creative teaching and active learning.

> *“We have the freedom to propose methodologies and supplement the course content. I know my colleagues do the same. We have discussed with some teachers—at various points—how we incorporate additional material that aligns with the overall direction without losing focus” (Teacher 2)*.

Another strength lies in the experience and academic background of the teaching staff. Although the course focuses on positive mental health, the instructors bring additional elements that enrich the subject matter and methodology. This reinforces active learning and creative teaching approaches—aspects that will be discussed later. Some interviewees note that they coherently integrate theoretical tools from psychology, social work, and anthropology regarding mental health.

Those who have taught the course over several semesters add that their experience allows them to enrich the content and teaching methods to reach students more effectively—especially given that most indicated the course material resonates with them both in their professional practice and on a personal level.

> *“You learn as you go; the course I teach is never exactly the same. I enjoy seeing how students respond to certain activities and thinking about how I can improve things—or reflecting on topics that I only truly grasped through the experience of teaching them. I find myself thinking, “Next time I teach this course, I’m going to change this aspect.” It’s about thinking in terms of how a course is built and evolves over time” (Teacher 3)*.

Regarding the course content, both instructors and students value the fact that the subject allows them to explore, discuss, and delve into everyday topics they might not otherwise pause to reflect upon. Students find the material relevant to the university context and the stage of life they are currently in. Overall, they are able to relate the topics to their own thoughts, feelings, and daily practices.

> *“The topics are well-suited for opening us up a bit and learning about the subject matter; after all, many of us hear about things like assertive communication or stress management—topics that often seem somewhat disconnected—without really focusing on researching them or finding ways to learn more or acquire those tools. This course consolidates all those important topics and presents them to you”. (Student, 5)*

The relevance of the topics is complemented by the implementation of methodologies that facilitate interactive processes between students and teachers. According to the students, priority was given to methodologies based on unconventional activities that allowed them to explore new forms of expression, self-awareness, and learning. Artistic and sensory activities stood out among these, fostering meaningful learning for the students.

### Participants in the focus groups stated

> *“Activities like the silhouette exercise, painting a mandala, or music therapy are things not commonly used—things you don’t do in everyday life—yet they lead you to see them as a viable option” (Student 3)*.

Regarding these class dynamics, both students and teachers highlighted the central role of the students and their active participation—grounded in the exploration of personal experiences related to the topics addressed each week—as a key strength. This dynamic stimulated the development of communication skills and autonomy in the learning process; it also progressively fostered relationships and an atmosphere of trust where students could express themselves freely and construct knowledge collectively. Focus group participants noted:

> *“I really liked that we were encouraged to be very participatory ourselves. I mean, the main focus was getting everyone involved; we carried out the activities ourselves (…)” (Focus Group 1)*.

For their part, the teachers identified changes in the students’ skills and attitudes. Improvements were seen in the ability to work in groups and speak publicly about their thoughts and feelings, alongside the opening of possibilities to build support networks with other students, the teacher, and other university departments. One teacher commented:

> *“The course content itself doesn’t change, but it does provide elements that help them realize they were facing a problem—and that they aren’t the only ones—while also allowing them to connect through empathy and create a support network among themselves” (Teacher 2)*.

The strength generated by the possibility of collective feeling and thinking in the classroom leads students to recognize it as a safe space—one where they can express themselves, but also where they can rely on others or offer support to them. Viewing the classroom as a safe space enabled students who struggled with social interaction and expressing their thoughts and emotions to be heard without fear of judgment, within a framework of mutual respect and validation. This environment fosters trust and emotional openness, facilitating learning from a relational perspective.

> *“I feel that everyone expressed themselves, and people didn’t judge you; instead, they understood you” (Student Focus Group 2)*.

Building trust allowed students to share deeply personal experiences—both verbally and through individual written work using a pedagogical tool known as a *bitácora* (or reflective journal), which was also valued as a safe space. They were required to work on this journal weekly, reflecting on their experiences in each class. They viewed it as a space where they could express themselves intellectually, emotionally, and creatively, confident that what they wrote would be treated with validation and respect and kept confidential by the instructor. They commented:

> *“(…) you can talk about a topic and know it won’t be discussed outside the class; when you write in the journal, the teacher isn’t going to go out and tell everyone about it (…)” (Student Focus Group 1)*.

Similarly, the opportunity to reflect on themselves and listen to others strengthened their ability to ask for help and build support networks involving instructors, students, and other university departments. Some instructors coordinated the course with various services or departments, either by having them present their offerings to the students or by referring specific cases for assistance. Referrals were made to the *Campus Diverso* program for gender-related issues, to University Student Services for information on subsidies or academic field trips, and to the Psychological Service to introduce students to available care pathways or initiate the support process. A teacher commented:

> *“There have been complex cases [that] come to light in the logbook. I have met with students individually to talk, and based on that, I recommend they begin the referral process with psychological services or social work. The logbook is often the first place where they reveal deeply troubling issues affecting them—even impacting their academic performance.” (Teacher P3)*

The instructors acknowledge that in this course, they fulfill the role of counselors—in addition to their teaching duties—by directly addressing certain student demands or needs and acting as mediators with other university services or departments. They are also aware that providing this support is challenging due to time constraints regarding student contact and the complexity of certain cases.

> *“(…) There are very complex cases, and simply activating referral pathways is not enough. (…) The challenge as an instructor is to be careful not to open wounds and leave them exposed, but rather to know how to provide support and guidance, ensuring the person gains access to a program or professional care.” (Teacher 2)*.

Students described various personal transformations resulting from their experience in the LS course. Self-reflection and self-awareness were recognized as tools for navigating aspects of their university and personal lives, particularly regarding emotional regulation and stress management.

> *“It helped me discover a part of myself I didn’t know existed. Or, more than anything, it helped me think about who I am” (Student – Focus Group 2)*.

> *“I feel the course really helped me understand my attitudes and emotions so I could manage them much better. And when I finished the course, I felt very satisfied because I achieved what I had wanted from the start.” (Student – Focus Group 2)*.

A positive impact on academic performance was also evident through the acquisition of study strategies and personal organization skills. Furthermore, from a physical health perspective, some students reported improvements in symptoms such as migraines and muscle pain.

> *“It really helped me get organized. It also helped with my study methods” (Student - Focus group 1)*

> *“It reduced things like my back pain and migraines. So, it was very beneficial for me” (Student – Focus group 2)*.

Finally, the importance of the course during the early semesters of undergraduate study was acknowledged, given its contribution to adapting to the university environment and to students’ personal lives:

> *“(…) I thought it was great that they included this in the first semester, because it gave us many tools for the semesters ahead—for managing stress and things like that, (…).” (Student – Focus group 2)*.

However, certain challenges were identified that may conflict with some of the aforementioned strengths. Some students held a negative predisposition due to the course title and its elective status; furthermore, the content and the highly emotional, experiential methodology posed obstacles for the instructors, as these factors influenced both the expected and unexpected outcomes of the course. For many students, such courses are viewed merely as a means to an end—a way to earn academic credits—without requiring significant effort or sparking genuine interest. Moreover, exploring and expressing aspects of emotion, personality, and character is far removed from their standard university training; indeed, doing so publicly is neither a common nor a comfortable experience for them.

> *“I have noticed that student resistance to these courses stems not only from viewing them as ‘filler’ but also from the discomfort they cause—a discomfort regarding the demands placed upon them and the difficulty involved, which they are often unwilling to consciously admit. It is not a matter of studying until a concept or formula is understood, but rather of working through topics and activities that evoke feelings. The notion persists that these topics are ‘alien’ to men” (Teacher 3)*.

The instructors identified several factors affecting the course’s delivery: class size, the diversity of ages and academic programs, students’ prior university experience, and expectations regarding the course content based on its title. Large classes hinder participatory activities, place a heavy burden on instructors—particularly regarding the exhaustive review of reflective journals—and limit the ability to identify and monitor cases requiring psychosocial support. They suggest group sizes of 25 to 30 students.

While student diversity is viewed as a positive asset, it also presents a challenge for instructors, as it involves dealing with segmentation based on academic program, age group, or other characteristics; this creates the dual challenge of fostering group cohesion while simultaneously addressing specific individual needs.

> *“The challenge for me was ensuring that the older ones could feel like part of it—and the younger ones too—and that the older ones could contribute to the younger ones’ process (…) You start looking beyond just the content and asking them: ‘What are you studying? Why? Are you in the program you want?’ and you begin to connect” (Teacher 2)*.

## Discussion

The findings are interpreted through a constructivist approach, which posits that knowledge is actively constructed rather than being a faithful replica of a pre-existing reality [24–25]. From this perspective, the cultural, economic, social, and political structures that condition both students and faculty are recognized as constituent levels of the educational process. This framework also incorporates the biographical experiences of participants and the emergent outcomes generated during the teaching-learning process [26].

At the intersection of social structures, life histories, and classroom interaction, faculty members act as mediators between students and culture to catalyze strategies promoting meaningful, interactive, and dynamic learning. Simultaneously, students participate actively in constructing their own knowledge through exploration, experimentation, and reflection. In this context, learning is viewed as an active process—“learning by doing“—where the body, mind, and emotions are engaged in action [27–30].

The following discussion is based on the perspectives of the interviewed students and faculty. While the sample did not include the entire student population, the data provide significant insights into the conditions of possibility and the achievements of the course regarding the strengthening of mental health and specific Life Skills (LS). Furthermore, the findings are influenced by the personal histories and emotional, cognitive, and attitudinal predispositions with which students enter the course, which affects their engagement and the intervention’s potential effects. The scope of the analyzed course must also consider structural factors such as session frequency: a three-hour weekly session over 16 weeks without subsequent continuity. Additionally, while participatory environments are designed to stimulate reflection and dialogue, these conditions cannot be unequivocally controlled by faculty or students, and their implementation does not invariably guarantee the desired emotional or psychological outcomes[31].

The analysis of findings is organized around three primary axes: 1) university education as a socialization process where faculty and students are conditioned by social structures yet retain margins of freedom and capacity for agency; 2) the classroom conditions that enable the teaching-learning process to impact LS and mental health; and 3) the specific positive transformations observed in students’ LS and mental health.

Based on these axes, the following hypothesis is proposed to guide the discussion and inform future research: in higher education contexts, flexible and alternative courses—distinct from technical-disciplinary training—enable active socialization processes that strengthen LS in university students, provided they occur under conditions of possibility where meaningful learning based on supportive relationships and dialogic, affective communication plays a central role.

Although recent studies rarely focus on courses explicitly designed for LS training, similarities are identified in the analysis of alternative disciplinary fields using flexible methodologies. There is consensus regarding the importance of elective courses in implicitly developing LS, which are fundamental for integral student development. These courses foster critical thinking and communication—essential tools for academic, personal, and professional life. Regarding practical and expressive methodologies, findings align with existing literature indicating that these strategies strengthen empathy, self-confidence, interpersonal relationships, personal development, and stress management, while facilitating adaptation to the university environment [20,31]. A literature review highlights a further point of convergence: the critical role of emotional mediation in teaching-learning processes [32].

In relation to the latter, research has identified correlations—not explored in the present study—between psychosocial well-being (self-esteem, life satisfaction, and perceived control) and academic performance [15]. Similarly, a link exists between active coping skills and the achievement of academic goals; students who utilize problem-solving strategies and seek social support demonstrate a greater capacity to overcome academic challenges and reach their educational objectives.

By articulating LS changes with the conditions of possibility within the framework of education as socialization, courses of this nature constitute novel educational environments. Despite having an established curricular structure, they are less predictable than traditional disciplinary subjects due to their high experiential and emotional load. As socialization spaces, they enable adaptation to new contexts while simultaneously fostering critical stances toward social norms, beliefs, or patterns [33] through students’ and faculty members’ lived experiences and classroom dialogue.

In this sense, courses where students and faculty act as protagonists—rather than external experts—facilitate the negotiation of the established social order alongside participants’ biographies during daily classroom interactions. This allows for the analysis and potential transformation of these structures, contrasting with subjects that prioritize instrumental knowledge. In these socialization and educational processes, the objective is not for students to memorize content or self-help formulas. Rather, the aim is for students to think, feel, and speak about themselves through the lens of each LS addressed in class, utilizing experiential and contextualized learning dynamics.

A key condition enabling these transformations is the horizontality of relationships between faculty and students, which allows for the reconfiguration of pyramidal authority structures. This recognizes the reciprocal feedback between participants as subjects with experiences and communicative capacity to produce collective knowledge [34]. A fundamental requirement for this type of relationship is the power of dialogue and communication in the classroom, utilizing both verbal and non-verbal language to express feelings, emotions, and needs [35].

This dialogue enabled faculty to recognize students’ specificities and extend class time to address needs through other university departments. Consequently, higher education institutions expand their formative functions to include tasks of prevention, early diagnosis, treatment, or referral [1]. These institutional networks broaden the socialization processes occurring in the classroom and provide essential support for student mental health.

However, these classroom relationships also face limitations, particularly regarding time constraints. All communication and social coexistence are process-oriented and do not occur immediately; they require intrinsic progress. Both students and faculty must gradually internalize the novel “codes” of the course to establish levels of communication and trust that transcend language and involve the body and emotions [35–37]. Therefore, time restrictions may prevent horizontal relationships from fully taking root for many students, especially when compounded by prior educational imaginaries.

Another vital condition is the centrality of subjectivities (interpretative frameworks, sensitivities, personality traits) and intersubjectivities. These are formed by the life histories and roles assumed by participants but also represent potentialities for transformation [38]. These subjectivities are enhanced when the curriculum focuses on relevant and pertinent themes that address the individual and social needs students face [39].

This capacity for self-reflection and dialogue is expressed through reflexivity in education, a condition required to transcend mechanical teaching-learning processes. In this regard, faculty members act as active and innovative agents who build strategies based on student profiles and the unique relationships woven within the classroom. For students, reflexivity emerges from the stimulus to think cognitively and emotionally about their own behaviors and those of others. Through language, emotional exploration, and creativity, students can reformulate their thinking to achieve transformations in LS and mental health [40]. As noted in the literature, reflective thinking allows for the transformation of practices and contributes to resignifying culture to respond to individual needs [41].

Finally, the findings provide evidence that the course had measurable effects on student mental health. It provided tools and enabled the development of knowledge, skills, and attitudes to navigate their realities both individually and through social bonds with others or institutions [31]. Specifically, it strengthened self-awareness, fostering the introspective capacity to identify needs and strengths while recognizing the possibility of change.

This stimulates autonomy in identifying and addressing problems, which facilitates help-seeking behaviors. This constitutes a salutogenic factor where interpersonal and social skills converge to build support networks that strengthen self-confidence, self-esteem, and the sense of belonging [42–43]. These coping strategies align with the WHO definition of LS as the ability to successfully manage the demands and challenges of daily life [31,44].

In conclusion, the participatory nature and the atmosphere of intimacy and trust generated in the classroom strengthened communication, listening, and empathy. These practices promote social interaction that transcends instrumental contact, fostering reciprocity and reinforcing the creation of support networks [45].

## Conclusions

The Life Skills course demonstrated a significant impact on student mental health. Key strengths of the strategy included a robustly structured, planned, and organized curriculum grounded in a Positive Mental Health framework. The implementation of specialized educational and didactic tools facilitated the achievement of course objectives and learning outcomes, with evaluation evidence indicating measurable progress in the development of ten core life skills among students.

The pedagogical approach transcends theoretical instruction by positioning the individual and collective analysis of student experiences as the central axis of learning. Furthermore, the creativity and expertise of the faculty expanded their roles from traditional content transmitters to facilitators of reflexive, affective, and experiential processes. This close pedagogical accompaniment enabled the identification of individual student needs and facilitated referrals to other university departments for specialized support.

Such courses represent a significant strength in higher education by offering experiential spaces that transcend technical-disciplinary boundaries. Based on horizontal educational relationships, a dialogic dynamic is promoted where students and faculty participate equitably to collectively foster an environment of respect, trust, and confidentiality. Consequently, the classroom is transformed into a safe environment for emotional expression and collective reflection, where mutual support and the recognition of others are integrated as essential components of the formative process.

A limitation of this study was the restricted diversity within the focus groups regarding student age, academic programs, and current semesters, which could have further enriched the findings. Additionally, while changes in Life Skills (LS) were identified through student focus groups and faculty interviews, these qualitative reports were not explored with the same precision as the quantitative component of the broader research project. These results underscore the necessity for future research to triangulate data from both methodological approaches to provide a more comprehensive analysis of the intervention’s effects.

## Data Availability

We agree to make the information in the document public once our manuscript is accepted for publication.

## Notes

### Competing Interest Statement

The authors have declared no competing interest.

### Author Declarations

The research was approved by the Health Research Ethics Committee, Faculty of Health, Universidad del Valle, under Approval Act No. 013-024.

